# Estimating the economic burden of stroke among informal caregivers in Australia using Productivity Adjusted Life Year

**DOI:** 10.1101/2024.11.21.24317753

**Authors:** Neha Das, Dieu Nguyen, Peter Lee, Suzanne Robinson, Lan Gao

## Abstract

**Background:** The Productivity Adjusted Life Year (PALY) metric is a novel measure for exploring the societal impact of health conditions on work productivity. Given that stroke is a major cause of DALYs (disability-adjusted life years) and the crucial role of caregivers in post-stroke care our study aims to quantify the PALY loss associated with stroke caregivers and the impact on their quality of life (QoL) by calculating quality-adjusted life years (QALYs).

**Method:** Dynamic life table modeling was used to simulate the follow-up of the Australian working-age population (aged 15–65 years) over 30 years of follow-up (2023-2052 inclusive). The model compared the Australian stroke informal carer group and a hypothetical cohort of non-carers. Key data was obtained from literature and publicly available data. The model was constructed using Microsoft Excel® (Microsoft Corporation, Redmond, WA, US). Labor force participation rates were applied to population estimates to arrive at the approximate number of full-time equivalent (FTE) which was then multiplied by the productivity index and years of life lived (YLL) to estimate PALYs and the utility values were multiplied by YLL to estimate QALYs.

**Results:** Over a 30-year period, an estimated 35,337 QALYs and 87,773 PALYs and AU$ 19.1 billion were lost due to the health and productivity burden attributed to the provision of informal care for stroke patients. On average, annual informal carer losses were 1,178 QALYs and 2,926 PALYS, corresponding to AU$ 636 million per year in productivity loss.

**Conclusion:** Our analyses demonstrate the health and economic consequences of providing care to stroke patients over the forthcoming decades and highlight the need for interventions to reduce the burden of stroke and its impact on caregivers.

## Introduction

Acute and chronic health conditions have a profound impact on the work productivity of millions of individuals worldwide. To assess the work-related effects of health conditions, factors such as absenteeism (days of work missed due to a condition), presenteeism (reduced productivity while working due to a condition), and premature exit from the workforce are often taken into account.^1^ Several validated tools are available to measure work impairment caused by health conditions, such as the Work Productivity and Activity Impairment (WPAI) tool.^2^ However, these measures typically assess productivity loss within a specific study sample over a short period and do not provide a comprehensive picture of productivity loss at a population level or over the course of an individual’s working life.^3^ The Productivity Adjusted Life Year (PALY) metric is a novel measure for addressing this knowledge gap by offering a broader perspective on the societal impact of health conditions on work productivity.^1^ To estimate the number of PALYs, the years of life lived (YLL) are multiplied by the productivity indices (which are conceptually similar to utility values) and the proportion of full-time equivalent (FTE) workers. Thus, the PALY collectively accounts for productivity loss due to absenteeism and presenteeism (based on the productivity index), labor force dropout (based on FTE data), and premature mortality (based on the risk of all-cause mortality and the resulting years of life lost), representing a more comprehensive measure for quantifying the productivity impact at the population level.^4^ PALYs are conceptually similar to quality-adjusted life years (QALY) but adjust years of life lived for impairment in productivity rather than impairment in quality of life. In other words, PALYs measure disease burden by quantifying the population-level impact of disease on productivity and the economy.^1^

Based on the 2021 Global Burden of Diseases (GDB) study,^5^ stroke was the third-leading cause of death and disability, contributing to 143 million DALYs (disability adjusted life years) (5.7% of total DALYs) in 2019.^5^ Moreover, up to 50% of stroke survivors are chronically disabled, and the risk of stroke recurrence over 5 years is considerable.^6,7^ In Australia, there were an estimated 27,428 stroke events, and 445,087 survivors of stroke in 2020.^8^ By 2050, without action, the number of strokes in Australia could increase to 50,600 and in such scenario, an estimated 819,900 Australians would be living with its impacts.^8^

Informal caregivers play a crucial role in post-stroke care^9^ (defined as individuals who provide care to those who need it within the context of an existing relationship, such as a family member, a friend, or a neighbor).^10^ The most significant improvements in stroke patients occur in the first few weeks post-stroke, with the majority of spontaneous recovery occurring within six months.^11^ However, many are left disabled with an impact on functional independence^11^ making caregiving an integral part of post-stroke recovery.^12^ The cost burden attributed to caregiving is considerable, even at 10 years post-stroke.^13^ If caregivers fall within the working age bracket, caring can impact their workplace productivity.^14^ Given the burden of stroke and the role of caregivers, it becomes imperative to understand the productivity impacts attributed to stroke in terms of the societal, population-level economic burden.

Moreover, studies have also demonstrated that there are negative impacts on the health and quality of life (QoL) of informal carers.^15–19^ Notably, there are statistically significant differences in mental (*p* = 0.003) and general health (*p* = 0.001) outcomes based on the Short Form 36 Questionnaire (SF36),^16^ as well as a significantly increased risk of chronic health issues such as high blood pressure (Odds ratio [OR]: 1.22) cardiovascular disease (OR: 1.29), diabetes (OR:1.43), asthma (OR: 1.49), and arthritis (OR:1.34)^15^ among carers as compared to non-carers.

Thus, our study aims to estimate the population-level productivity loss of informal caregiving in stroke by quantifying the PALY loss associated with stroke caregivers. Additionally, this study will explore the impact on QoL of stroke caregivers by calculating QALYs.

## Methodology

### Economic Model

A dynamic life-table model with a 30-year time horizon was constructed, using 2023 as the base year. A 30-year time horizon is chosen, as the majority of informal carers in Australia are aged ≥ 35 years^10^ and the average age to access a pension in Australia (66 years).^20^ This timeframe aligns with the average age of the cohort and allows for the analysis of potential labor force impacts on absenteeism/presenteeism. The model compared two cohorts: the Australian stroke informal carer group (‘Informal carer’) and a hypothetical cohort of non-carers (‘Non-carer’). The model included the working-age (aged 15–65 years) population. The working age population was defined as that used by the Australian Institute of Health and Welfare (AIHW) which is used to report labour force definition data.^21^ Deaths were assumed to occur halfway through each annual cycle. The population of informal carers was updated in each cycle by considering the new cohort of informal carers arising out of change in the stroke population and the death of informal carers. The model was constructed using Microsoft Excel® (Microsoft Corporation, Redmond, WA, US). Key data was obtained from literature, and publicly available data reported by the Australian Bureau of Statistics (ABS), AIHW, and Survey of Disability Ageing and Carers (SDAC). Outcomes accrued beyond the first year of the modeled simulation were discounted at 5.0% per annum as recommended by the Pharmaceutical Benefits Advisory Committee (PBAC).^22^

### Model Inputs

Key model inputs are presented in Table 1.

**Table 1:**
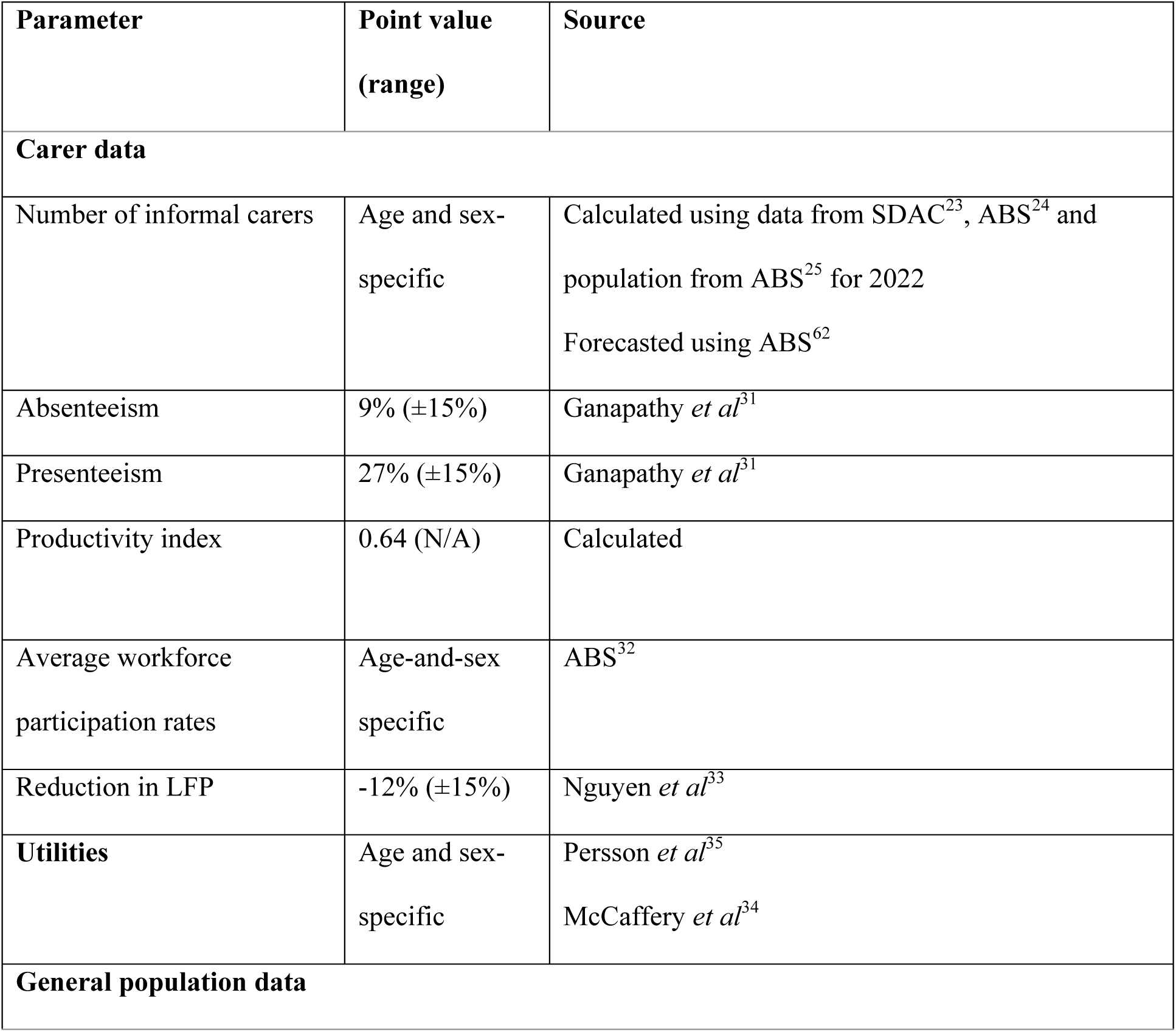
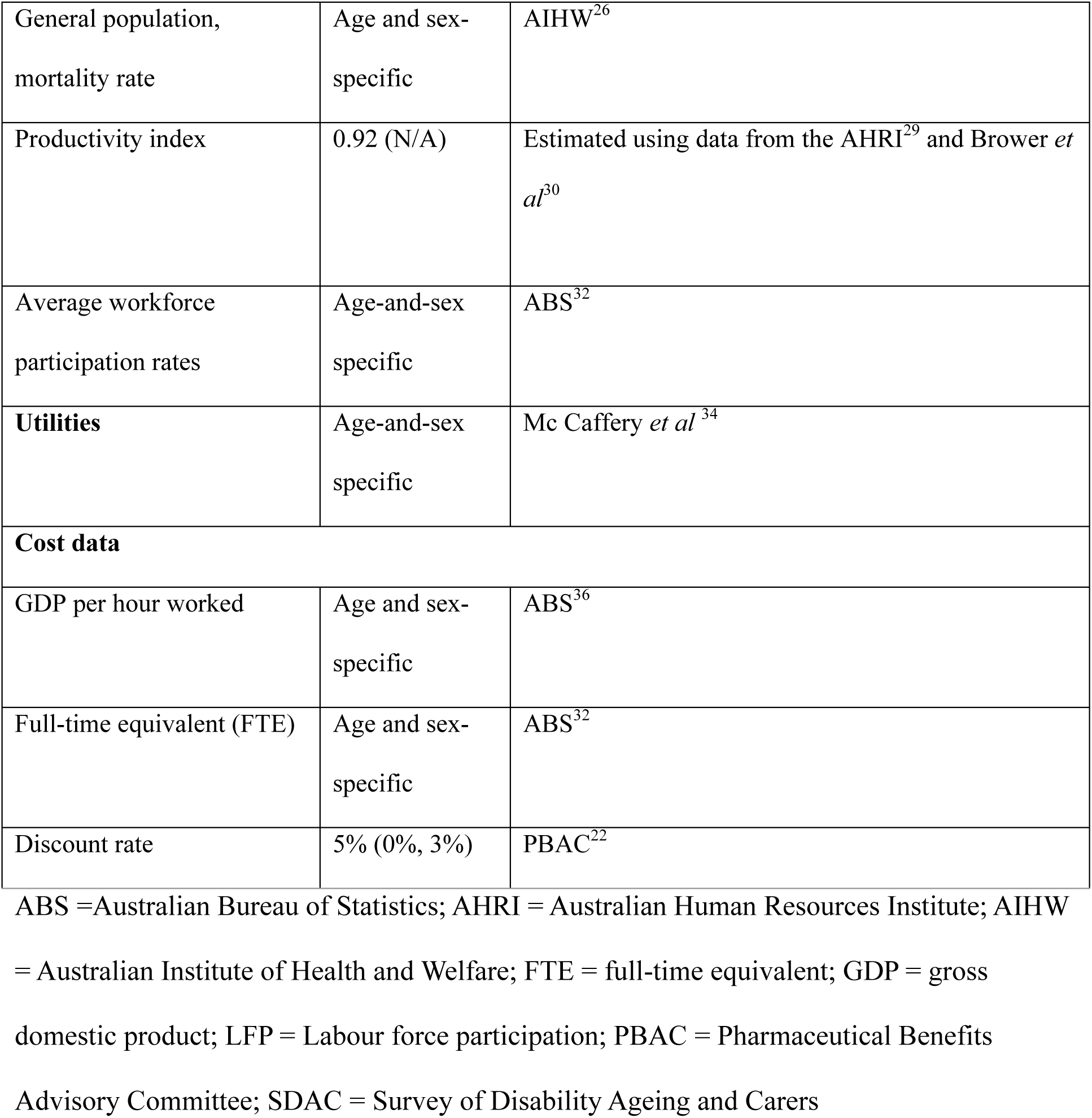
Key model inputs.

#### Deriving the informal carer cohort

Data from the ABS and SADC were used to inform the number of informal caregivers. The SDAC was used to inform the number of caregivers where the main condition of the main recipient of care was stroke.^23^ The SDAC provides this data only for primary carers. As such, this analysis captured PALY loss due to informal carers who were primary caregivers; that is, outcomes for informal carers who were not primary caregivers were not captured in the economic model.

First, to estimate the number of informal carers by age and sex, the number of primary carers for patients with stroke in 2022 (N = 26,100) was weighted by the proportional distribution of all primary carers in Australia by 10-year age group (ranging from persons aged 15 to 24 to 75 and over) and sex from the SDAC^23^ and ABS^24^ (Supplement Tables S1 and S2 in the Supplement). This was subsequently weighted by Australian population data from the ABS to quantify the number of informal carers for patients with stroke by age (single years) and sex (male/female) in 2022 (Supplement Table S3).^25^

The 2022 population represented the model population in the year prior to the baseline year of 2023 as the latest carer data was available for 2022. The model was evolved from 2023 onwards, across each age stratum by considering the entry (new carers for prevalent stroke patients) and exit (informal carer mortality). To estimate the number of informal carers for patients with stroke over a 30-year period, the prevalent population of stroke patients over time across male/female sexes was extrapolated using prevalence data for the years 2003, 2009, 2012, 2015, and 2018 from the SADC^23^ using linear regression modeling (Supplement Table S4). The linear regression model was run on 2003, 2009, 2012, 2015, and 2018 data and 2022 data was used to validate the model. Next, male and female carers as a percentage of the total stroke population were calculated using the data from 2022. This percentage was applied to the number of estimated stroke patients to calculate the total number of male and female carers in each year from 2023 onwards. Subsequently, the age-and-sex stratified proportion of informal carers in 2022 was applied to the predicted carers in each annual cycle to estimate the number of informal carers for patients with stroke across age (single-year) and sex (male/female) strata over a 30-year time horizon (2023-2053 inclusive) (Supplement Table S5).

To estimate the number of deaths occurring in each annual cycle to inform the exit of non-carer/informal carers from the model, the population in each age stratum was multiplied by the age-specific risk of all-cause mortality. This was estimated using mortality data from the AIHW throughout the modeled time horizon (30 years).^26^ The mortality risk between carers versus non-carers was assumed to be equivalent on the basis that there is limited evidence regarding differences in mortality risk between them.^27^

### Modeled outcomes

#### Informal carer productivity burden

The model estimated the productivity burden attributed to providing informal care for patients with stroke using PALYs, which captures health-related productivity loss arising from absenteeism or presenteeism. Productivity indices were estimated by dividing the number of days worked within a year without absenteeism or presenteeism, with the maximum number of working days in a year.^28^ On average, absenteeism among the Australian population was 8.8 days per work year based on a report from the Australian Human Resources Institute (AHRI);^29^ average presenteeism (10.7 days per work year) was sourced from a study by Brower *et al* conducted in the Netherlands.^30^ The rate of absenteeism (9%) and presenteeism (27%) per week among informal carers were sourced from Ganapathy *et al.,*^31^ which explored the productivity burden attributed to providing informal care for patients with stroke in the USA. In Australia, the maximum number of working days in a year is 240, which reflects that the population, in general, has four weeks of annual leave per year and works five days per week.^28^ This corresponds to productivity indices of 0.92 and 0.64 for non-carers and carers, respectively.

Labor force participation rates (both full-time and part-time where the hours were converted to FTE assuming 40 full-time working hours) for the working age (15–65 years) population were sourced from the ABS for 2023, stratified by sex and age in single years.^32^ To reflect the impact of providing informal care for stroke patients on labor force participation rates, we assumed that the rate of non-participation in the labor force for informal carers was 12 percentage points lower than the rate of non-participation for non-carers.^33^ Labor force participation rates for the working age (15–65 years) population were applied to population estimates to arrive at the approximate number of FTE workers for each age stratum in the carer cohort and, for the general population. The FTE was multiplied by the productivity index and YLL in the model to estimate PALYs.

#### Health-related Quality-of-life burden

Utility values reflect a person’s health-related quality-of-life (HRQoL) based on physical and mental health domains.^34^ They are estimated using generic, multi-attribute utility instruments (MAUIs), and lie on a scale from 0 (representing dead) to 1 (full health), with negative values representing health states considered worse than death.^34^ Normative Australian age and sex-specific utilities were applied to non-carers in the economic model.^34^ The impact of informal caregiving on HRQoL was estimated using data from a study that compared HRQoL between spouses of dependent and independent stroke survivors, and controls in the Netherlands.^35^ The utility weight ratio between spouses of dependent stroke patients (mean utility weight: 0.69), and the control population which included spouses of the normal population (mean utility weight: 0.77) was applied to normative Australian population utilities to estimate the HRQoL burden attributed to providing informal care for patients with stroke^34^ (Supplement Table S6). These utility values were multiplied by the estimated YLL to estimate QALYs in the model for informal carers and non-carers.

#### Cost of lost productivity

All costs are presented as 2022 Australian Dollars (AU$). The cost attributed to lost productivity was estimated by assigning a cost value for each PALY. The unit cost for a PALY in any given year was based on the GDP per FTE worker for the corresponding year (2023-2052 inclusive). Data pertaining to GDP per hour worked were sourced from the ABS^36^ and used to estimate the GDP per FTE for a given year based on a 40-hour work week and a maximum of 48 weeks worked per working year. For example, the GDP per FTE worker in 2022 was estimated to be $192,000 based on a GDP per hour worked of AU$100 x 40 hours x 48 weeks. The temporal trend in GDP per hour worked from 1975 to 2023 was used to project the GDP per FTE worker for the informal carer/non-carer population over the modeled period (2023 – 2053 using univariable linear regression model to extrapolate GDP per hour worked over time and R^2^ as goodness of fit). For reference, the unit cost of a PALY in 2023 was estimated to be AU$193,522, increasing to AU$256,458 in 2052.

### Sensitivity analysis

A variety of deterministic sensitivity analyses (DSA) were performed through varying key model parameters. These were varied individually while other variables were maintained at base case values to explore the impact of key parameters on modeled outcomes. Key parameters assessed include presenteeism and absenteeism, number of carers, stroke prevalence, and labor force participation. In addition, we evaluated the impact of varying the discount rate. The lower and upper bounds for the variables for sensitivity analysis are reported in Table 2 and in Supplement S7.

**Table 2:** Data for sensitivity analysis.

| **Parameter** | **Range** | **Source** | **Distribution** |
| --- | --- | --- | --- |
| Discount rate | 3 - 5% | <sup>63</sup> | Uniform |
| Change in informal carer (base case no change) | 0 - 2.7% | <sup>9</sup> | Uniform |
| Change in stroke prevalence | -1.13% - 2.07% | <sup>5</sup> | Uniform |
| Change in LFP* | ±15% | <sup>28,38</sup> | Uniform |
| Change in absenteeism/presenteeism** | ±15% | <sup>28,38</sup> | Uniform |

LFP = labour force participation
Uniform distribution was used using Lee 2023,<sup>28</sup> Satyana 2020,<sup>37</sup> and Hartmanis 2021<sup>38</sup>

A probabilistic sensitivity analysis (PSA) was undertaken to examine the joint uncertainty of the key model input parameters, such as cost, quality of life, and productivity. The PSA was performed by running 1,000 iterations of Monte Carlo simulations. The respective distributions for the variables were selected as per Briggs et al, which is in line with previous studies that performed economic evaluations with PALYs.^28,37,38^ It was assumed that parameter distributions were independent (i.e. the change in one parameter does not correlate with the change in another parameter).

## Results

The results of the base-case economic analysis are presented in Figures 1 and 2 below, and in Supplement Tables S8 and S9.

**Figure 1:**
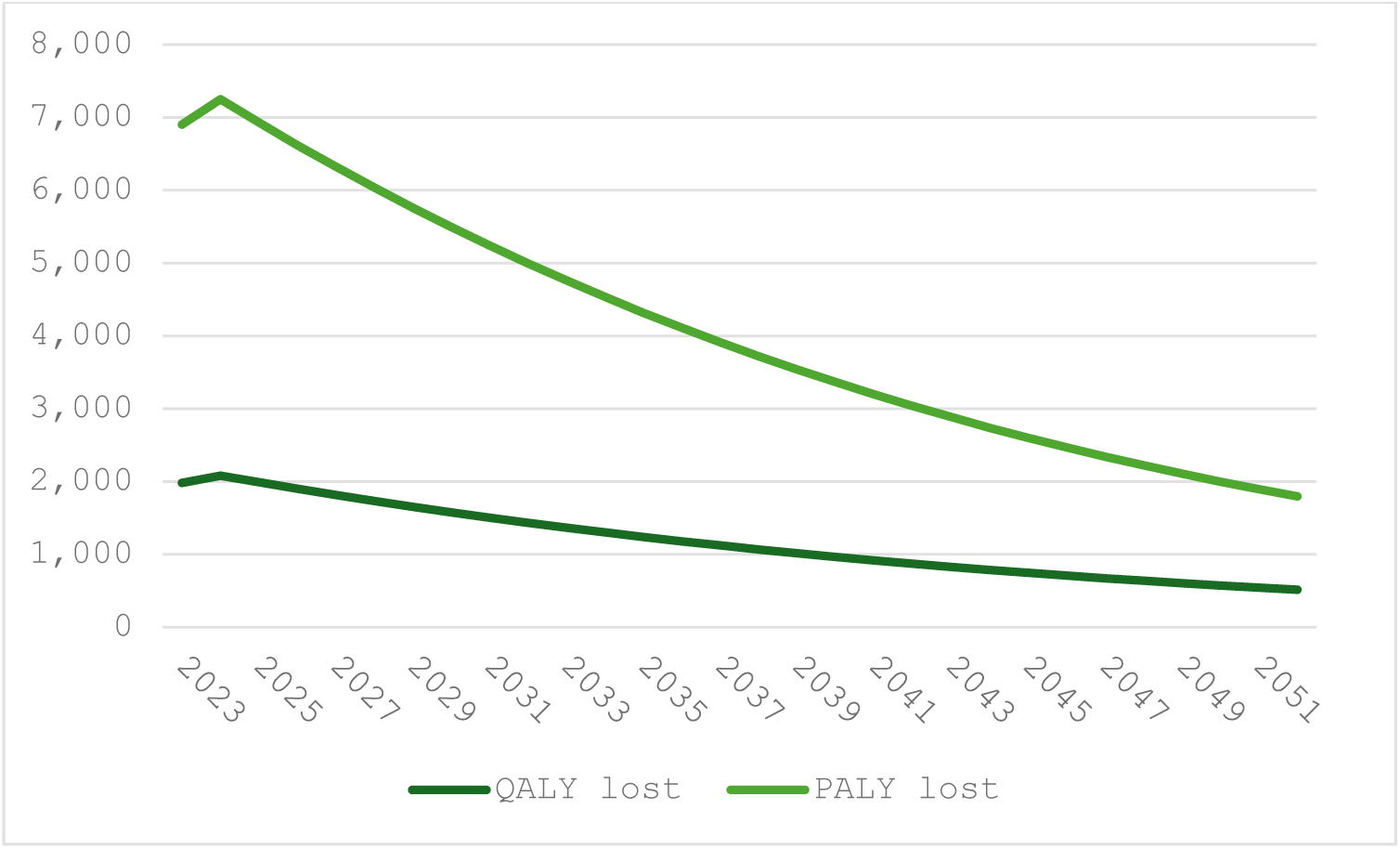
Results of the base-case analysis (PALY/QALY differences) PALY = productivity-adjusted life year; QALY = quality-adjusted life year

**Figure 2:**
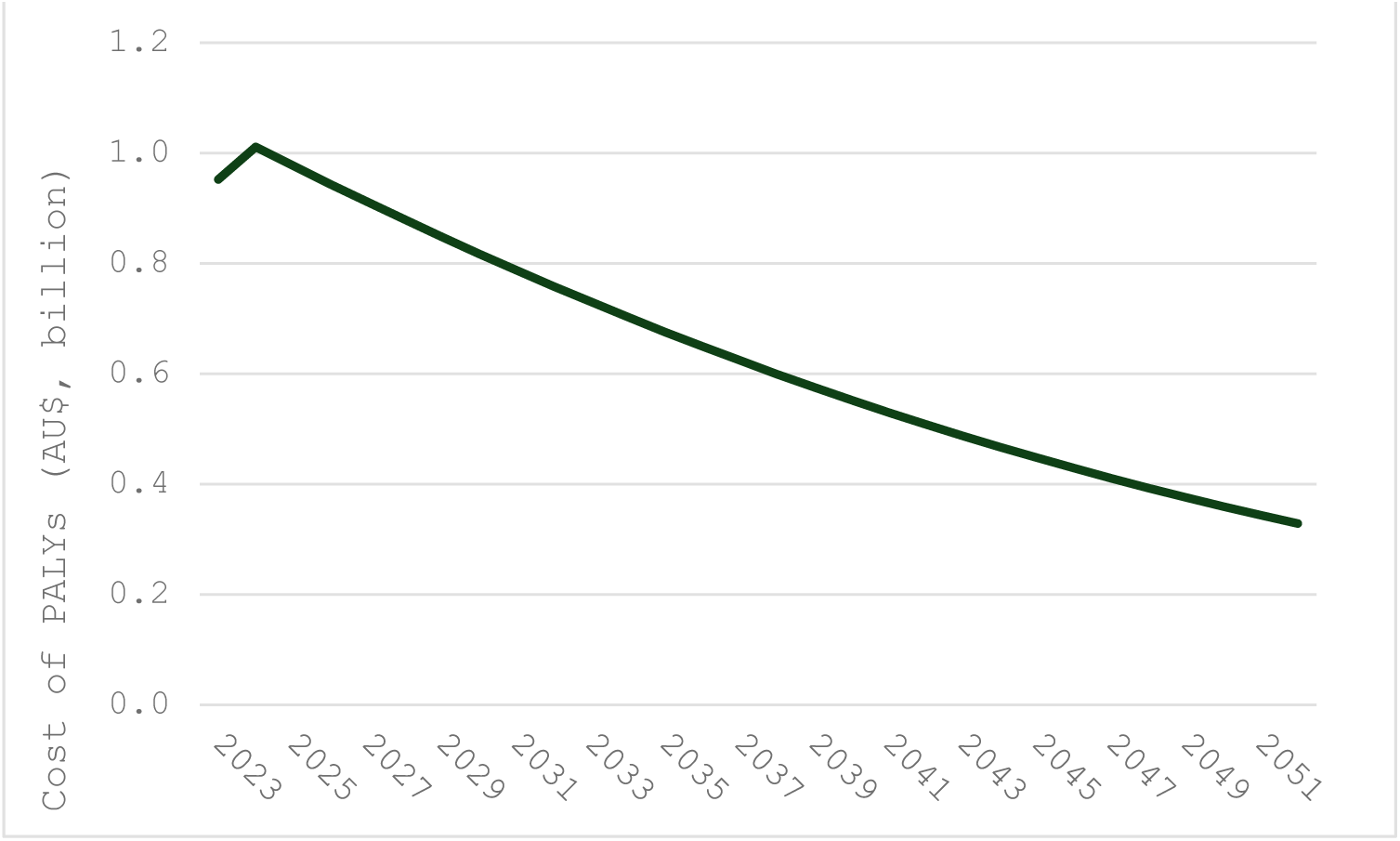
Difference in PALY costs over time. AU$ = Australian dollars; PALY = productivity-adjusted life year

Over a 30-year period (2023 to 2052 inclusive), an estimated 35,337 QALYs and 87,773 PALYs were lost due to the HRQoL and productivity burden attributed to the provision of informal care for stroke patients. Assuming the cost of each PALY increased over time, the total cost attributed to lost productivity among informal carers for patients with stroke over a 30-year period was AU$ 19.1 billion.

On average, annual informal carer losses were 1,178 QALYs and 2,926 PALYs, corresponding to AU$ 636 million (M) per year in productivity loss. Upon stratification by sex, the average QALY and PALY loss due to caregiving among males was 331 QALYs, 966 PALYs, and AU$ 210 M per year. The total HRQoL and productivity burden for males was 9,922 QALYs, 28,986 PALYs, and AU$ 6.3 billion over a 30-year period. Among female informal carers, the average HRQoL and productivity losses were 847 QALYs and 1,960 PALYs respectively, corresponding to AU$ 426 M per year in productivity losses. The total QALY and PALY losses among female caregivers were 25,415 QALYs, and 58,787 PALYs, corresponding to AU$ 12.8 billion.

### Sensitivity analysis

Table 3 presents the results of DSA, as well as the results from 95% confidence interval values from the PSA for QALYs and PALYs lost, and the cost of PALY lost. Key model drivers include the discount rate assumed, the time horizon considered, changes in the number of informal carers over time, and changes in absenteeism and presenteeism. Reducing the time horizon to 15 years reduced the estimated number of lost QALYs (23,882 versus 35,337) and PALYs (59,310 versus 87,773), as well as the associated loss in productivity costs (AU$ 12.3 billion versus AU$ 19.1 billion). An increase in the number of informal carers by 2.7% led to an increase in PALY loss and cost of PALY loss by 33,421 and AU$ 7.8 billion, respectively from baseline. The impact of a hypothetical scenario in which the stroke prevalence increased by 2.07% annually, led to an increase in PALY loss of 1,774 and an increase in the cost of PALY losses of A$ 387 M from baseline. Contrary to this, a 1.13% annual decrease in stroke prevalence led to a reduction in PALY loss and cost of PALY loss of 968 and A$ 211 M respectively from baseline.

**Table 3:** Results of the deterministic and probabilistic sensitivity analyses.

| **Parameter** | **QALY lost** | **PALY lost** | **Cost of PALY lost** |
| --- | --- | --- | --- |
| Base-case | **35,377** | **87,773** | **\$19,080,824,765** |
| Discount rate (base case 5%) |  |  |  |
| 3% | 44,210 | 109,818 | \$24,188,604,042 |
| Change in informal carer (base case no change) |  |  |  |
| 2.7% | 48,789 | 121,194 | \$26,831,580,898 |
| Change in stroke prevalence |  |  |  |
| Lower (-1.13%) | 34,947 | 86,805 | \$18,869,718,147 |
| Upper (2.07%) | 36,051 | 89,547 | \$19,467,542,198 |
| Reduction in LFP* |  |  |  |
| Smaller reduction (12%*0.85) | - | 84,025 | \$18,266,156,890 |
| Greater reduction (12% *1.15)) | - | 91,521 | \$19,895,492,639 |
| Change in absenteeism* |  |  |  |
| Lower 15% | - | 85,085 | \$18,496,566,014 |
| Upper 15% | - | 90,460 | \$19,665,083,515 |
| Change in presenteeism* |  |  |  |

| Parameter | QALY lost | PALY lost | Cost of PALY lost |
| --- | --- | --- | --- |
| Lower 15% | - | 79,767 | \$17,340,185,750 |
| Upper 15% | - | 95,904 | \$20,848,193,780 |
| 15-year time horizon (2023-2037 inclusive) | 23,882 | 59,310 | \$ 12,272,811,620 |
| Results of PSA |  |  |  |
| 95% CI |  |  |  |
| Lower bound | 47,009 | 88,591 | 19,570,024,778 |
| Upper bound | 47,183 | 92,135 | 20,356,175,992 |

\*Change to variables applied only to the carer cohort
CI: Confidence interval; DSA: Deterministic sensitivity analysis; GDP Gross domestic product; LFP Labor force participation; PALY productivity-adjusted life year, PSA: Probabilistic sensitivity analysis; QALY quality-adjusted life years

## Discussion

The present study fills an important gap in the literature by quantifying the productivity burden borne by caregivers of stroke patients. We estimated that the provision of informal care for patients with stroke would lead to an estimated 35,337 QALYs and 87,773 PALYs lost, corresponding to AU$ 19 billion in lost productivity over a 30-year period (2023 – 2052 inclusive). Few recent studies have explored the impact on caregivers or families due to stroke. A recent report by The Stroke Foundation highlights that the lifetime cost of the impact on household productivity (defined as the hours of time spent performing domestic activities, childcare activities, adult care activities, and voluntary work) was A$ 3 billion and the lifetime cost of impact on the informal caregiver (defined as a person most closely involved in helping the person with stroke to live at home) was another A$ 4 billion.^39^ Another study by Tan et al, 2022 reported the non-medical cost (including informal care) of stroke among young adults as 537 million over 30 years.^40^ Moreover, of the Australians of working age who were missing from the labor force and giving care to a relative or friend living with them, stroke and hypertension were among the five most common conditions among the care recipients.^41^ As such, our findings provide justification for efforts that reduce the productivity burden attributed to the provision of informal care for patients with stroke.

Previous research estimated the productivity loss associated with stroke patients^42–45^ but few studies have estimated lost productivity among caregivers of stroke patients. One such study found that employed caregivers of stroke patients faced a productivity loss of US$10,020 (AU$ 15,235 using a conversion rate of 1.52) per year (versus AU$ 27,156 from the present study).^31^ This difference is attributed to the inherent difference between productivity loss and PALY loss and consequently, the calculation methodology in our study is different from that used by Ganapathy *et. al*.^31^ While Ganapathy *et. al*. estimated productivity loss per patient (in terms of hours/days) and calculated the cost by attributing labor cost value to it, our study includes absenteeism, presenteeism, and workforce participation rates, and applied GDP values to calculate the productivity of a caregiver. We further calculated the loss as compared to a cohort of the same population had they not been carers. A similar approach was adopted by another study which calculated the PALY of parental and control groups for those caring for children with Angelman’s syndrome comparing the PALYs lived between these cohorts.^38^ The PALY lost over 10 years was 495 and the PALY loss cost was A$ 45.3 million.^38^ The per-parent PALY loss was 1.50^38^ in this study, versus 0.125 per carer in our study. While the approach is the same, this variation is due to the number of carers which differ among these studies.

Our study highlights that there is a gender disparity among caregivers, with a higher number of female caregivers than males. The AIHW reports that 72% of all primary carers are females (in 2018).^10^ This is also reflected in previous studies, which have shown that females are more likely to provide informal care.^46–53^ Furthermore, female caregivers report experiencing a higher caregiving burden than males.^46,50,54^ Similarly, in our study, the PALY loss was higher among females than males. The disproportionate impact of caregiving has important policy implications. Efforts to support informal caregivers should consider the needs of female caregivers, including access to respite care and employment assistance.

Globally, there has been a decline in the age-standardised stroke incidence, prevalence, death, and DALY rates from 1990-2019.^5^ In Australia, there were decreases in prevalent cases (29%), deaths (58%), and DALYs (58%) from 1990 to 2019^5^ based on the GBD 2019 study and in age-standardized rate of stroke events from 2000 onwards as reported by the AIHW.^55^ However, despite the decline, stroke continues to contribute to 4.9% of all deaths and 20% of cardiovascular disease deaths in 2021^56^ and has an economic impact of 9 billion per year in Australia.^39^ These findings underscore the importance of stroke preventive and management strategies. As an example, a recent report by the Stroke Foundation reports that achieving 30/60/90 National Stroke Targets could lead to almost $26 million in savings over a lifetime (for the strokes that occur in any one year).^39^ Notably, although the Stroke Foundation Report includes the cost of informal care, it excludes the cost of caregivers’ PALY loss, and therefore might not indicate the considerable productivity cost impact among them.

Caring requirements can have a significant impact on a caregiver’s employment status, with many forced to reduce their working hours or withdraw from the labor force.^9^ Based on ABS data on Australian carers, primary carers are less likely to be in labor force (58.8%) as compared to non-carers (81.5%).^57^ Additionally, female carers are less likely to be employed (63.2%) than male carers (71.8%).^57^ Given the impact of caring on employment, policies that help to increase caregivers’ participation in the workforce or support them in employment may ultimately mitigate the PALY burden attributed to informal care. Some strategies, such as the Victorian Carer Strategy 2018-22 have been established to support caregivers.^58^ The priorities include better health and well-being for caregivers, support in school, study, and work environments, support and services that meet their needs, help with financial stress, and recognition and respect for caregivers.^58^ Other examples include government services such as the Carer Gateway programme which provides online skill courses, support packages (services or equipment to help with education, help with cooking, cleaning, shopping, and transport), and emergency respite care for carers.^59^ Additionally, the Carer-Inclusive Workplace Initiative was created following recommendations from the Australian Government’s Jobs and Skills Summit to foster a more inclusive, supportive workplace for Australians who provide unpaid care in the community.^59^ The initiatives reflect a commitment to support caregivers’ employment. However, other factors that influence a caregiver’s decision to remain within the workforce and work productivity, such as family circumstances, financial resources, expectations regarding informal caregiving and employment, and employers’ views on work commitment, require a comprehensive understanding of the caregivers’ situation to develop robust policies which will improve productivity and employment.

The demand for informal carers in Australia is estimated to grow significantly over the next decade, with a rise of 23% from 1.25 million to 1.54 million by 2030.^9^ Furthermore, the supply of informal carers will not be proportional to the needs.^9^ Our sensitivity analysis revealed that the change in informal carer number had the highest impact on PALY loss. The productivity impact due to the informal carers can be improved through implementing strategies to minimize stroke disability and thus dependence on caregivers as well as implementing policies to support and accommodate informal caregivers in employment. Legal policies such as the Equal Opportunity Act 2010 enable carers to manage their work and caring responsibilities effectively. However, there are other aspects such as whether carer employees can discuss the difficulties they are facing with employers and whether employers in turn can provide arrangements such as flexible work hours, part-time work, or paid/unpaid care leave hours that may be required by the carers.^41^ Some provisions are available in Australia to support caregivers, such as income support and respite care.^10^ However, there are also limitations as well, for example, regarding aged residential respite care, Carer’s Australia 2018 Report notes that the key barriers for carers included unaffordability, ineligibility for respite care, high needs of the person receiving care, and low availability.^60^ Thus, a comprehensive understanding of patients’ and caregivers’ needs and designing solutions accordingly could help informal carers and eventually may improve both their productivity and their quality of life.

We acknowledge certain limitations of this study. Firstly, while studies have reported that caregiving is associated with poorer mental health,^16^ and an increased risk of chronic health issues such as cardiovascular disease, diabetes, asthma, arthritis, and osteoporosis^15^ as compared to non-carers, other studies have reported no statistically significant difference in HRQoL among the two groups.^61^ Thus, we did not explore the cost associated with health conditions experienced by informal caregivers. Secondly, in lieu of suitable data, we adopted an aggregate approach to derive the age and gender-stratified number of informal caregivers. Moreover, we made certain assumptions regarding absenteeism, presenteeism, and HRQoL among informal carers due to the unavailability of comprehensive data. While this may not accurately reflect the Australian settings and contribute to uncertainty around the outcomes, the results of our sensitivity analyses supported the robustness of our findings.

## Conclusion

Informal carers play an important role in managing patients with chronic conditions and 10.8% of all Australians are informal carers.^9^ Our analyses demonstrate the health and economic consequences of providing care to stroke patients over the forthcoming decades and highlight the need for interventions to reduce the burden of stroke as well as the impact on caregivers.

## Data Availability

The data used in the analysis is included in the supplement.

## Acknowledgments

Not applicable

## Sources of Funding

This study was conducted as a part of PhD program for which Deakin University provided scholarship.

## Disclosures

Not applicable

## Notes

### Competing Interest Statement

The authors have declared no competing interest.

